# Intimate Partner Violence Experiences among Men Who Have Sex in Hanoi, Vietnam

**DOI:** 10.1101/2024.12.01.24318278

**Authors:** Loc Quang Pham, Patrick S. Sullivan, Amanda K. Gilmore, Ameeta S. Kalokhe, Thanh Cong Nguyen, Khanh Duc Nguyen, Hao Thi Minh Bui, Le Minh Giang

## Abstract

**Purpose:** Intimate partner violence (IPV) victimization is a pressing issue among men who have sex with men (MSM) and has profound health implications. This study aims to estimate the prevalence of IPV and identify factors associated with its occurrence among MSM in Hanoi, Vietnam

**Methods:** A cross-sectional study was conducted from March to May 2023 at an MSM-friendly sexual health clinic in Hanoi. Participants completed a self-administered questionnaire on a tablet after screening for eligibility. IPV victimization was assessed using a 21-item scale (the IPV-GBM scale). Logistic regression models were used to identify factors associated with IPV victimization in the past 12 months.

**Results:** Among 309 respondents, the mean age was 28 and half of participants reported using PrEP for 12 months or more. Two-thirds reported IPV victimization in the past 12 months (67%). Emotional IPV (47%) was most prevalent, followed by monitoring (39%), controlling (19%), and physical/sexual IPV (15%). Participants who were reported experiencing IPV in the past 12 months were more likely to be aged ≤ 24 years (aOR = 1.98, 95% CI: 1.14, 3.44), to report preferring the insertive sexual role (aOR = 2.02, 95% CI: 1.10, 3.70), or to report having more sexual partners (2-5 partners vs ≤ 1 partner: aOR = 2.02, 95% CI: 1.10, 3.70; ≥ 5 partners vs. ≤ 1 partner: aOR = 2.02, 95% CI: 1.10, 3.70).

**Conclusions:** IPV victimization among MSM on PrEP in Hanoi is highly prevalent. Tailored interventions are needed to address these vulnerabilities and promote safer behaviors and healthier relationships. Efforts should prioritize younger MSM. Further research is necessary to explore IPV dynamics across diverse settings.

## Introduction

Intimate partner violence (IPV) among men who have sex with men (MSM) is a pervasive health problem and is particularly relevant in the context of HIV prevention efforts.^1^ Intimate partner violence (IPV) refers “to any behavior within an intimate relationship that causes physical, psychological or sexual harm to those in the relationship,” including physical, sexual, economic, and emotional violence.^2^ IPV is common among sexual minority groups. A meta-analysis of 26 studies including more than 39,000 sexual minority people (SMP) in the United States found that SMP were 3.8 times more likely to experience sexual abuse compared to the general population.^3^ Another recently published meta-analysis of 52 studies including 32,048 MSM reported that the pooled prevalence of IPV victimization was 33%.^4^

Experiencing IPV is associated with significant short– and long-term health consequence.^5^ IPV is linked to an increase in suicidal ideation^6,7^, and homicide^8^. MSM suffering from IPV are more likely to engage in substance use and to experience depressive symptoms.^1,9,10^. MSM experiencing IPV are also more likely to engage in high-risk sexual behaviors, such as in unprotected sex, transactional sex and then have higher risk of acquiring HIV.^1,10–12^ Furthermore, MSM living with HIV and experiencing IPV have significantly worse HIV outcomes and engagement in care.^13^

Studies assessing violence among MSM in low– and middle-income countries (LMICs) remain limited.^4^ A study of 278 female sex workers, MSM, and transgender women in five countries in Latin America and the Caribbean reported that nearly all participants experienced some form of violence.^14^ Another study among young MSM in Myanmar reported 21%-25% of the participants experienced sexual violence.^15^ In Vietnam, a 2016 survey among 202 MSM found that 14.4% had experienced lifetime sexual violence, using a single self-reported item.^16^ Furthermore, prior research often lacked validated, multidimensional IPV instruments, leading to potential underestimation of IPV prevalence and its profile and associated factors.^2,4^

This study addresses these gaps by utilizing the validated IPV-GBM scale^17^ to comprehensively assess the prevalence and correlates of IPV among MSM in Hanoi, Vietnam.

## Methods

### Study overview

A cross-sectional study was conducted between March 27 and May 14, 2023, at an MSM-friendly sexual health clinic in Hanoi, Vietnam. Study participants were MSM receiving oral daily PrEP. Eligible participants self-administered a structured questionnaire using a tablet in a private room after providing informed consent. Ethical approval for the study was obtained from the Institutional Review Board at Hanoi Medical University.

### Study site

Hanoi has the second-highest estimated number of MSM among 64 provinces in Vietnam, with at least 30,000 MSM.^18^ HIV prevalence among MSM in Hanoi is estimated at 11.2%.^19^ The Sexual Health Promotion (SHP) clinic, established in 2013, is a trusted resource for the LGBT+ community, offering services such as HIV post-exposure prophylaxis, PrEP, and sexually transmitted infection (STI) testing and treatment. Currently, the clinic has provided PrEP services to approximately 2,000 users, of whom 60–70% use daily PrEP and the remainder use event-driven PrEP.

### Participants

Eligible participants were 18 years old or above, male at birth, had been taking oral daily PrEP for at least 30 days, and had at least one male sex partner or male lover in the past year. A sample size of 310 was determined based on an anticipated IPV prevalence of 60%^16,17^, with a precision of 10% and an estimated population size of MSM 180,000.^20^ To enroll participants, we screened all MSM visiting the clinic during the study period. Eligible but non-consenting individuals were asked to provide basic demographic and behavioral data, such as year of birth, months on PrEP, educational levels, employment, and having male sex partners or male lovers in the past year.

### Data collection

Participants completed a self-administered questionnaire on a tablet. Prior to starting, a research assistant explained key terms (e.g., defining “partner” as inclusive of regular and casual relationships and of sexual or emotional relationship) and instructed on how to use the tablet. Assistance was available throughout the process to address technical or comprehension issues. Participants received 150,000 VND (equivalent to 7.5 USD) for their time and effort. Surveys took approximately 30–35 minutes to complete.

### Measures

#### Intimate partner violence

The experience of IPV was assessed using the IPV-GBM scale.^17^ The scale was developed in English to be particularly used among gay and bisexual men and validated in the US.^17^ This 21-item scale includes five IPV domains: Physical and sexual IPV (6 items), monitoring behaviors (5 items), controlling behaviors (4 items), emotional IPV (3 items), and HIV-related IPV (3 items). Each item in the scale has five choices: “Never happened,” “1 time in the past year,” “2 times in the past year,” “3-5 times in the past year,” and “More than 5 times in the past year”. The scale underwent a rigorous five-step cultural adaptation process, including translation and cognitive interviews, to ensure validity for Vietnamese MSM.^21^

### Potential Correlates of IPV

#### Social support

Levels of social support were measured by an eight-item instrument modified from a nineteen-item Social Support Survey instrument.^22,23^ The modified scale measures two dimensions – tangible support (4 items) and emotional support (4 items). Respondents were asked “How often is each of the following kinds of support available to you if you need it?” (e.g. someone you can count on, to confide in, to love and make you feel wanted). Each item was rated from “none of the time,” to “all of the time,” with a score ranging from 1 to 5, respectively. Scores were computed to range from 1 to 100, with higher scores indicating greater support.

#### Sexual histories

The data included sexual preferences including whether they preferred penile-anal sex sexual role, the number of sex partners (in general, casual sex partner), condom use in anal sex, group sex, and sexualized drug use (e.g., methamphetamine or Viagra) in the past 12 months.

#### Background characteristics of the participants

Variables included age, highest educational levels, employment, marital status, personal monthly income, housing condition, sexual orientation, and duration of PrEP use.

### Data analysis

The primary outcome was any IPV victimization in the past 12 month. We focused past 12 months because it was an indication of intervention. It was coded as “1” for the participants with any experience of any 21 items of the IPV-GBM scale in the past 12 months and “0” for participants without any experience of any 21 items of the scale in the past 12 months. Similarly, participants experienced physical or sexual IPV, monitoring behaviors, controlling behaviors, emotional IPV, or HIV-related IPV if they experience any of the items belonging to corresponding form of IPV. Experience of IPV victimization was reported by frequency and percentage. Standardized Cronbach’s alpha was calculated to assess internal validity for the overall scale and its subdomains.

Socio-demographic and behavioral characteristics of the study participants were summarized by mean and standard deviation for continuous variables (e.g., age, social support) or by frequency and percentage (e.g., education, sexual orientation). Differences between participants with and without any IPV victimization were assessed by Chi-squared tests for categorical variables or by Student’s T-tests for continuous variables. Multivariable logistic regression models were used to identify factors associated with any IPV victimization. Variables were included in the model if their associations for association with IPV had p-values <0.2 in the univariate analysis. Adjusted odds ratios (aOR) and their respective 95% confidence intervals were reported. Model goodness-of-fit was evaluated using Hosmer-Lemeshow tests. Statistical analyses were performed using STATA version 18 (Stata Corporation, College Station, TX, USA), and a p-value < .05 was considered as statistically significant.

## Results

During the study period, 741 clients visited the SHP clinic, of whom 679 (92%) were using PrEP. Among these 679 PrEP users, 396 (58%) were eligible to participate in the study, and 310 (78%) of those eligible consented and completed the questionnaire (Supplement Table 1). Comparisons between participants who agreed and who refused to participate in the study showed that older and employed people were less likely to participate in the study (Supplement Table 2). One participant was excluded from the analysis due to event-driven PrEP usage, resulting in a final analysis sample of 309 participants (Figure 1).

**Figure 1.**
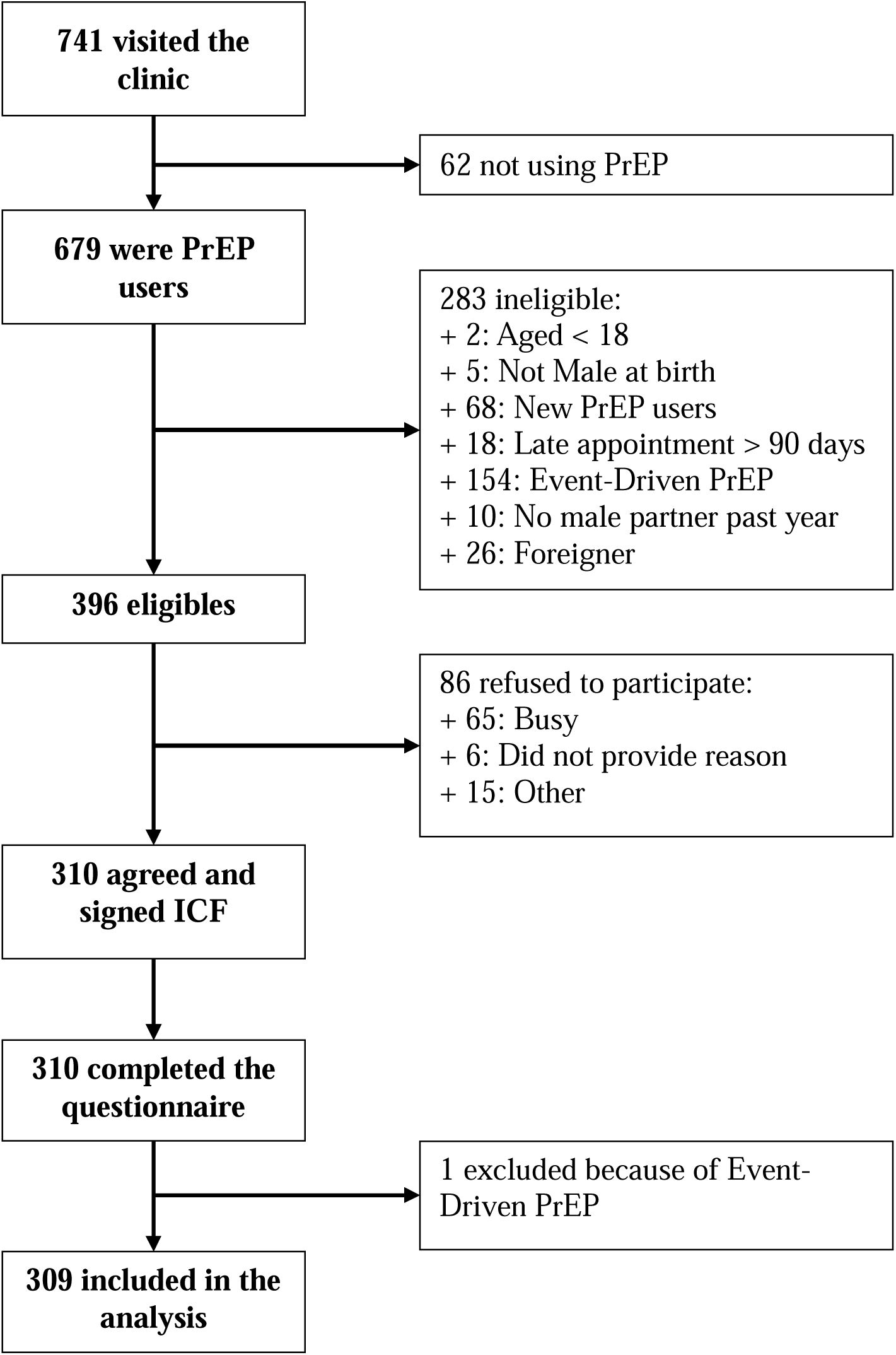
Study recruitment chart from March 27^th^ to May 14^th^ (7 weeks)

The mean age of the study participants was 28 years, with 36% aged 24 or younger. Most participant self-identified as gay (60%) and 17% reported experiencing housing insecurity within the past 12 months. The majority (55%) reported having 2–5 sexual partners in the past year. Most (53%) participants reported inconsistent condom use during anal sex in the past 12 months and 66% reported the use of sexualized drugs in the past 12 months. Approximately half (51%) of participants had been using PrEP for 12 months or longer (Table 1).

**Table 1.**
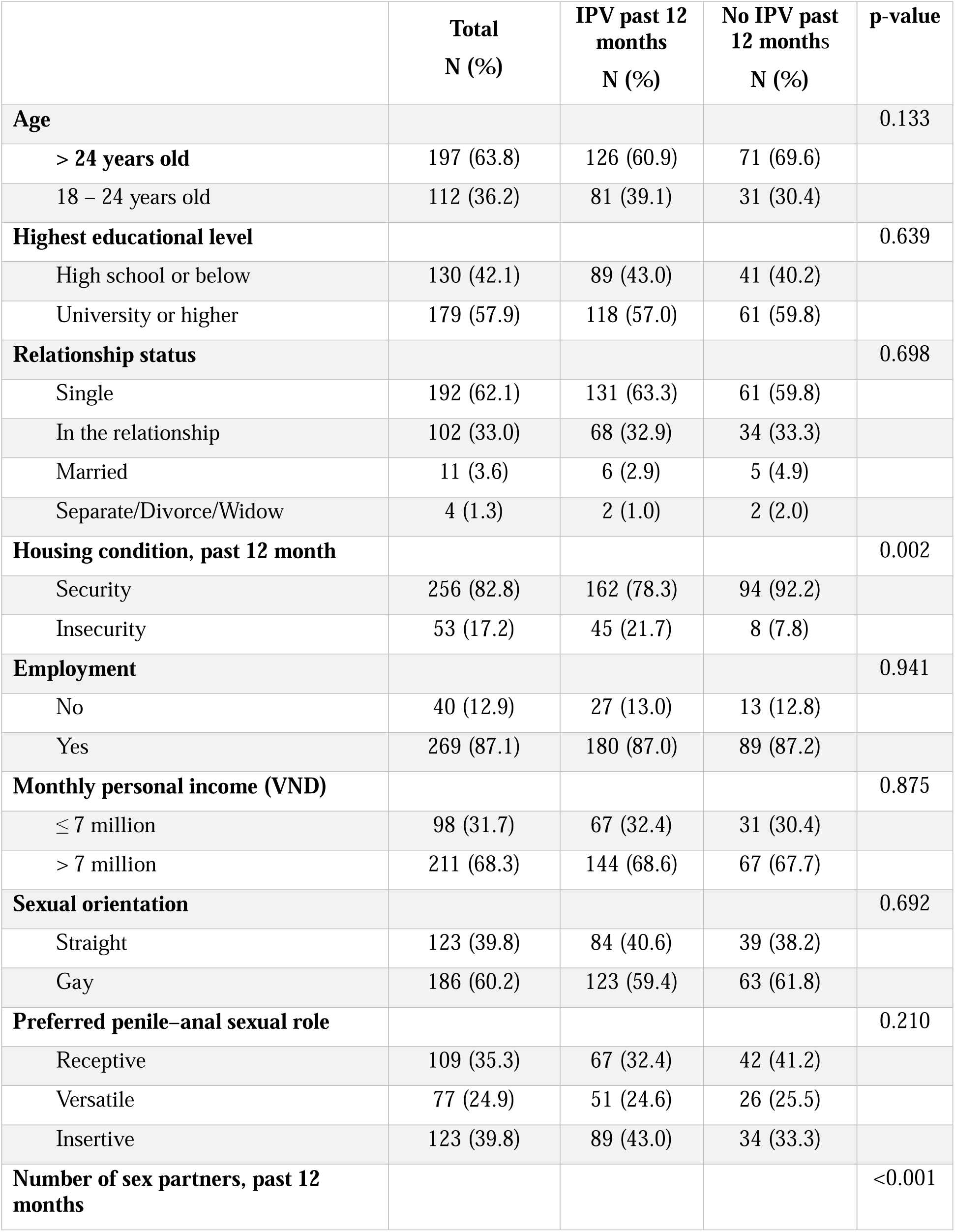
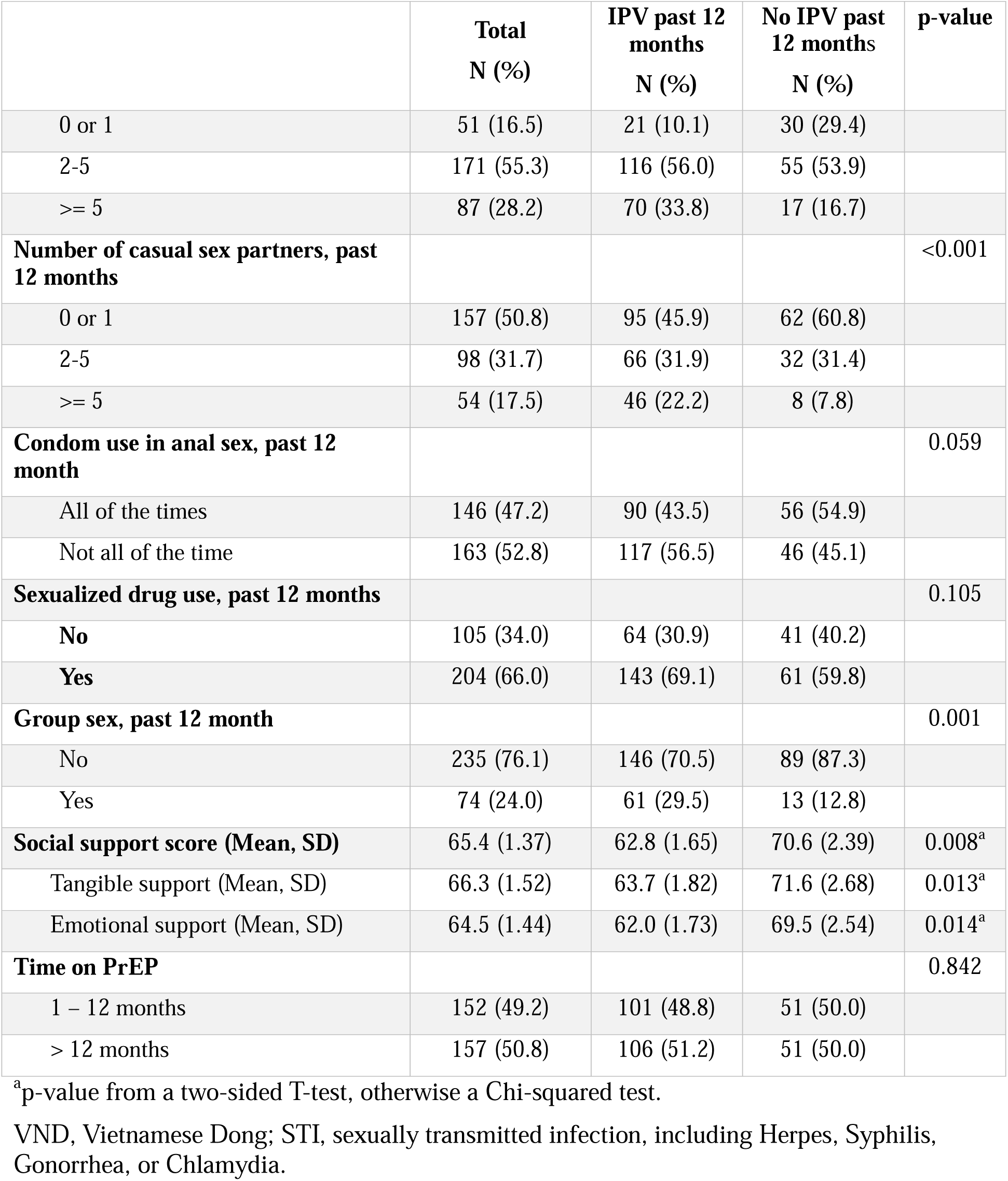
Demographic and behavioral characteristics of men who have sex with men using PrEP by experience of IPV in the past 12 months, Hanoi, Vietnam, 2023. (n = 309)

Among 309 participants, 67% experienced one or more forms of IPV in the past 12 months, and 68% experienced lifetime IPV. The prevalance of each form of IPV victimization in the past 12 months and in lifetime were similar. Emotional IPV was the most prevalent form (47% in the past year and 49% lifetime), followed by monitoring behaviors (39% and 40%), controlling behaviors (19% and 20%), and physical or sexual IPV (15% and 22%). HIV-related IPV victimization was 12% last year as compared to 20% lifetime prevalence. Most standardized Cronbach alphas were >0.7, indicating moderate to good reliability, excepting for controlling behaviors in the past 12 months (Cronbach’s alpha = 0.56) (Table 2).

**Table 2.**
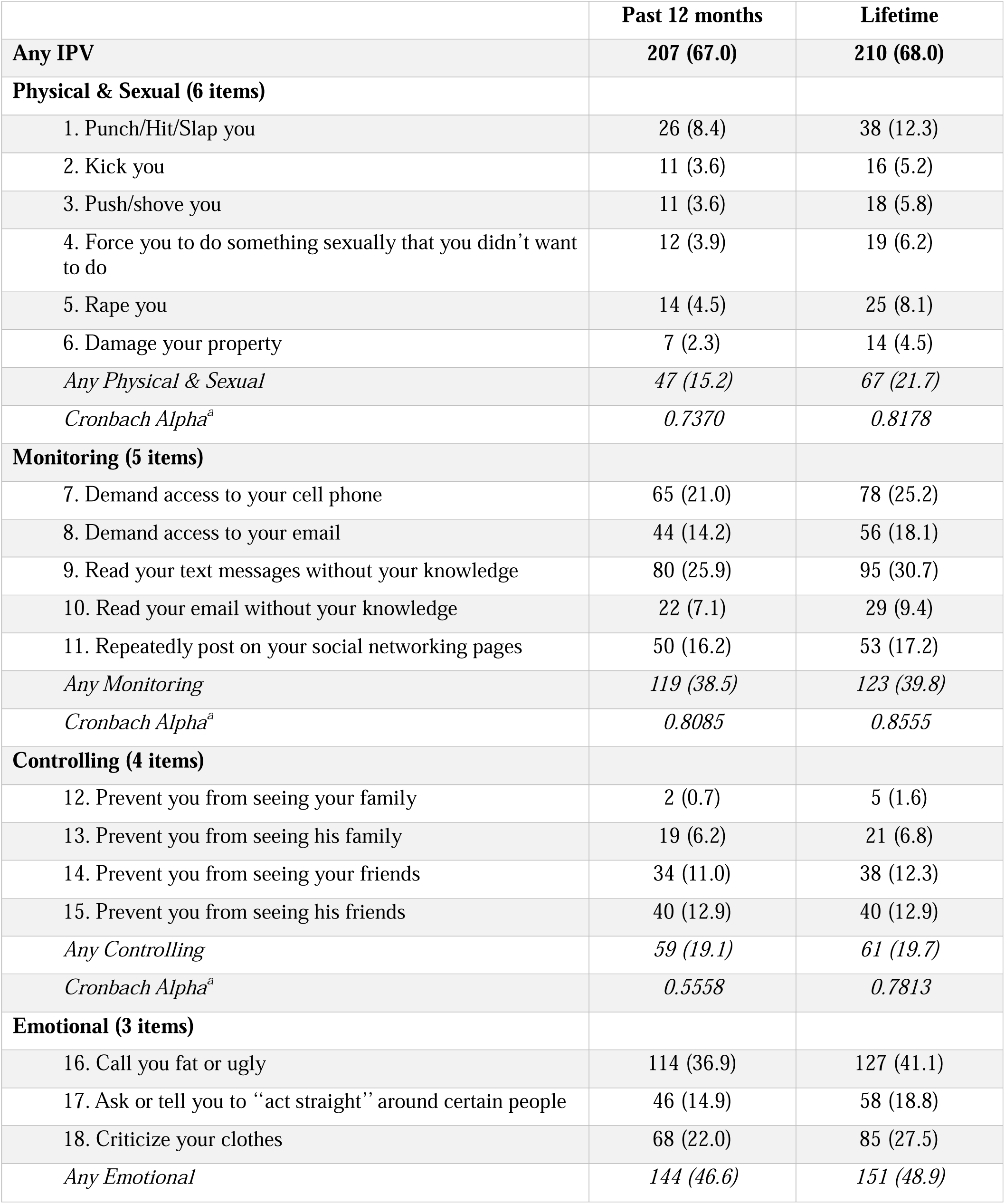
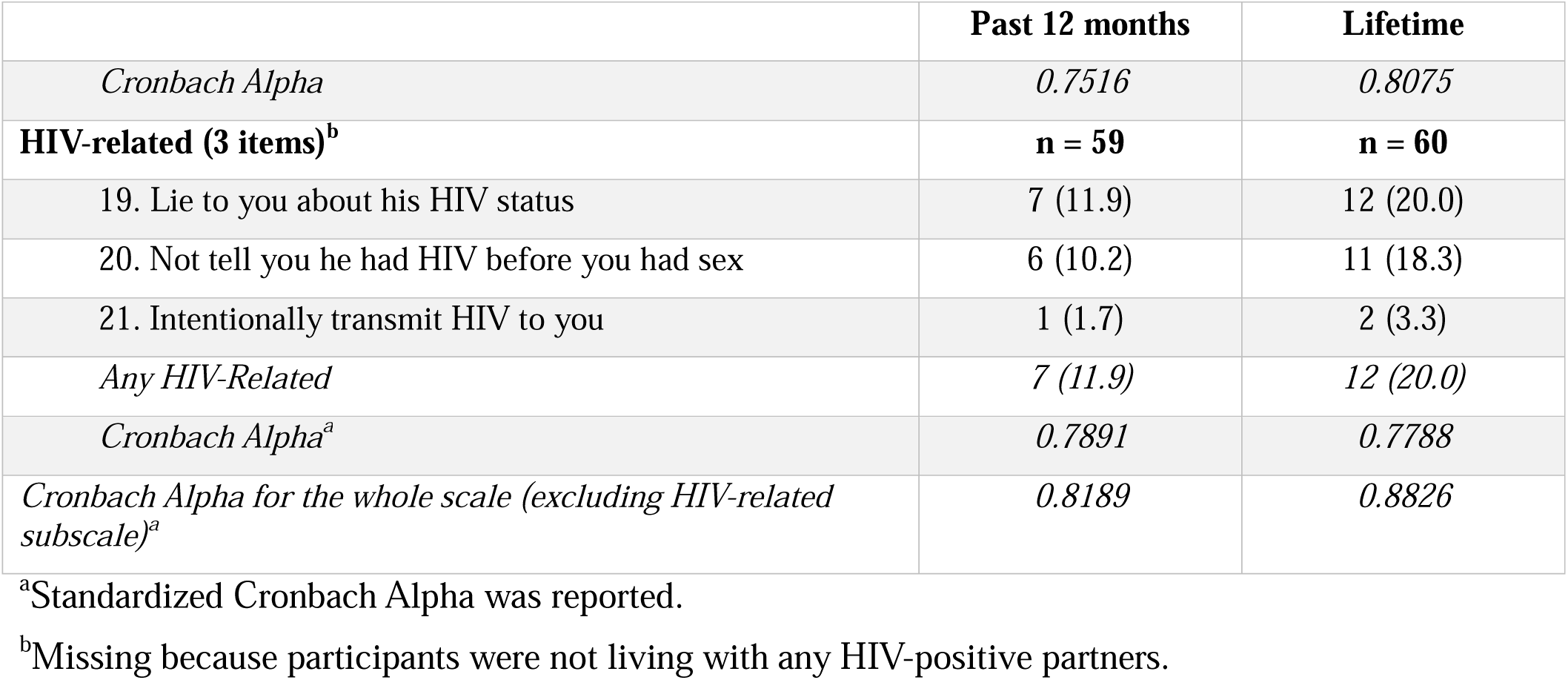
Percentage of intimate partner violence victimization among men who have sex with men using PrEP at a clinic in Hanoi, Vietnam, 2023. (n = 309)

Participants who reported housing insecurity had a three-fold higher proportion of experiencing any IPV victimization than participants who had a regular place to stay (22% vs. 8%, p-value 0.002). MSM with more sexual partners in the past 12 months were also more likely to report IPV. Additionally, MSM participating in group sex in the past 12 months were twice as likely to report last-year IPV victimization as compared to those not participating in group sex (30% and 13%, p-value 0.001). Lower levels of social support were observed among those who experienced IPV compared to those who did not (Table 1).

The multivariable logistic model suggested that participants aged ≤ 24 years were nearly twice as likely to experience any IPV victimization than those aged > 24 years (aOR: 1.98, 95%CI: 1.14, 3.44). Compared to participants who preferred receptive sexual roles, those who preferred insertive sexual roles were more likely to experience any IPV victimization (aOR: 2.02, 95%CI: 1.10, 3.70). A higher number of sex partners in the last year was positively associated with experience of any IPV victimization (2-5 sex partners aOR: 2.48, 95%CI: 1.26, 4.89; >=5 sex partners aOR: 3.37, 95%CI: 1.42, 7.99) (Table 3). Neither housing insecurity nor group sex participation was statistically associated with IPV in adjusted analyses. The p-value from Hosmer-Lemeshow test (p-value = 0.99) indicated the model fitted well.

**Table 3.** Multivariable logistics regression of experiencing IPV past 12 months on associated factors among men who have sex with men, Hanoi, Vietnam, 2023. (n = 309)

|  | Adjusted OR (95% CI) | p-value |
| --- | --- | --- |
| **Age group** |  |  |
| > 24 years old | 1 |  |
| 18 – 24 years old | 1.97 (1.12, 3.45) | 0.019 |
| **Housing condition, past 12 month** |  |  |
| Security | 1 |  |
| Insecurity | 2.00 (0.85, 4.70) | 0.112 |
| **Preferred penile–anal sexual role** |  |  |
| Receptive | 1 |  |
| Versatile | 1.24 (0.64, 2.39) | 0.531 |
| Insertive | 2.01 (1.10, 3.69) | 0.024 |
| **Number of sex partners, past 12 months** |  |  |
| 0 or 1 | 1 |  |
| 2-5 | 2.53 (1.29, 4.99) | 0.007 |
| >= 5 | 3.44 (1.45, 8.16) | 0.005 |
| **Condom use in anal sex, past 12 month** |  |  |
| All of the times | 1 |  |
| Not all of the time | 1.34 (0.79, 2.25) | 0.277 |
| **Sexualized drug use, past 12 months** |  |  |
| No | 1 |  |
| Yes | 1.26 (0.73, 2.16) | 0.407 |
| **Group sex, past 12 month** |  |  |
| No | 1 |  |
| Yes | 1.79 (0.84, 3.79) | 0.130 |
| **Social support score** | 0.99 (0.98, 1.00) | 0.070 |

IPV, intimate partner violence; OR, odds ratio; CI, confidence interval
The number of sex partners in the past 12 months was selected instead of the number of casual sex partners because of better performane, tested by Likelihood ratio test.

## Discussion

This study is among the few reporting the current circumstances of IPV victimization among Vietnamese men who have sex with men (MSM). It highlights that the prevalence of intimate partner violence (IPV) victimization among MSM using PrEP was high – about two-thirds of men reported experiencing IPV in the past 12 months. This prevalence of IPV victimization among our study participants exceeds that reported in studies in the US^24–26^, China^27,28^, and a previous study in Vietnam.^16^ Differences may stem from different recruitment methods. For example, prior studies recruited participants via online advertisements^24^ or from gay-friendly venues^25^, while our study recruited participants from a health clinic. Moreover, varying recall periods (e.g., 6 months in Sharma et al., 2021)^26^ and the use of fewer items to measure IPV in some studies^16,27,28^ may contribute to these discrepancies.

To our knowledge, this is the first study to adapt and use the IPV-GBM scale in Vietnam. Despite minor modifications following linguistic adaptation guidelines^21^, the scale retained consistency with its original form. We observed a similar pattern of different forms of IPV: the highest prevalence was emotional IPV, followed by monitoring, controlling, and physical/sexual IPV.^9,29^ Because the scale is novel for use in Vietnamese MSM, further research is needed to validate this scale within Vietnamese MSM populations and to explore short-form adaptations and optimal thresholds for intervention.

Consistent with previous studies, younger participants were more likely to report IPV in the last 12 months.^24,27^ Young MSM may lack experience in navigating interpersonal conflicts or ensuring their safety in relationships. Socioeconomic disparities and power imbalances in relationships with older partners may further heighten their vulnerability.^26,30^ Additionally, IPV exposure increases risks for HIV and STIs^10–12^, underscoring the need for targeted interventions for young MSM.

An important finding from this study – the positive association between the number of sexual partners in the last 12 months and IPV – is well-established in studies among women^31^ but has been uncommonly reported in studies among MSM. The mechanism could be complex. Different types of partners might be associated with different forms of IPV. For example, both regular partners and transactional sex partners were more frequently the perpetrators of IPV than casual partners.^9,27^ A more parsimonious explanation of this finding might be that having more partners means exposing oneself more frequently to partners capable of inflicting IPV. Establishing and adhering to sexual agreements in intimate relationships is crucial, as research has shown that men are more likely to experience IPV when their partners believe they have violated such agreements.^24,26^ Notably, having more than two sexual partners has been identified as an independent risk factor for both STIs^32^, and HIV infections.^33,34^ Therefore, promoting skills such as effective condom negotiation, developing and maintaining sexual agreements, and undergoing routine testing for HIV and STIs presents an opportunity to empower couples to address health concerns. These strategies may not only reduce IPV but also mitigate its associated consequences, including the risk of HIV infection.

The association of preference for an insertive role with a higher prevalence of IPV victimization is an important finding that, however, conflicts with the literature. One study suggested that MSM who had receptive anal sex during 30 days before the interview were more likely to experience any lifetime IPV.^35^ Qualitative studies are needed to contextualize this finding among Vietnamese MSM.

The study findings should be interpreted in light of some limitations. First, our study was conducted in a sexual health clinic in Hanoi; this may limit generalizability to broader MSM populations, especially those outside Hanoi or without healthcare engagement. However, the clinic is the setting where interventions will be implemented, and a large number of MSM already access to the clinic, so data generated from this clinic-based sample will inform the future development of clinic-based interventions against violence. Further research should engage a larger sample size and involve MSM at different clinics and practice types in other provinces. Second, we could not avoid the natural limitation of a cross-sectional design, in which reverse causation is possible. For example, in our study, condom use, or social support can be a consequence of IPV victimization, instead of a risk factor for it.

## Conclusions

This study underscores the urgent need for policies and interventions to reduce IPV among MSM in Vietnam. The study found the high prevalence of IPV victimization among MSM attending PrEP services in a sexual health clinic in Vietnam. Younger age, preference for an insertive sexual role, and higher number of sex partners were positively associated with IPV victimization in the past 12 months. Qualitative data should be collected to provide context and to provide more information about the circumstances and perceptions of these incidents, which guide the development of violent prevention intervention. Interventions to mitigate IPV should focus on screening to identify victims, providing more support for youth, and promoting safe behaviors and healthy relationship skills among and should integrate into PrEP/ARV settings where a large number of MSM already have access to.

## Authorship contribution

**Loc Quang Pham**: Conceptualization (lead), Methodology (equal), Data collection (lead), Formal analysis (lead), Original draft (lead), Review and editing (lead). Patrick S. Sullivan: Conceptualization (supporting), Methodology (equal), Original draft (supporting), Review and editing (supporting). **Amanda K. Gilmore**: Conceptualization (supporting), Methodology (equal), Supervision (equal), Original draft (supporting), Review and editing (supporting). **Ameeta S. Kalokhe**: Conceptualization (supporting), Methodology (equal), Supervision (equal), Original draft (supporting), Review and editing (supporting). **Thanh Cong Nguyen**: Data collection (supporting), Review and editing (supporting). **Khanh Duc Nguyen**: Data collection (supporting), Review and editing (supporting). **Hao Thi Minh Bui**: Data collection (supporting), Review and editing (supporting). **Le Minh Giang**: Conceptualization (supporting), Methodology (equal), Supervision (equal), Original draft (supporting), Review and editing (supporting).

## Data Availability

All data produced in the present study are be available upon reasonable request to the authors.

## Acknowledgments

The research reported in this manuscript was supported by Fogarty International Center under a training grant “Consortium for Violence Prevention Research, Implementation, and Training for Excellence” [D43TW012188]. The content is solely the responsibility of the authors and does not necessarily represent the views of the funding agency. The authors gratefully acknowledge the project team members in Vietnam and the US for their contributions to this study.

---

**Supplement Table 1.** Reasons for ineligibilities of clients who visit SHP clinic, Hanoi Vietnam, 2023.

|  | **Frequency** |
| --- | --- |
| Clients came to SHP | 741 |
| PrEP clients | 679 |
| In-eligible clients | 283 |
| Age < 18 | 2 |
| Not Male | 5 |
| M0 clients | 68 |
| Late > 90 days | 18 |
| Event-driven PrEP | 154 |
| No partner in the past year | 10 |
| Not invited | 26 |
| Eligible clients | 396 |
| Refuse to participate | 86 |
| Agree and sign ICF | 310 |
| Complete the questionnaire | 310 |

**Supplement Table 2.** The comparison between participants who participated and did not participate in the study.

|  | **Refuse to participate**<br>**(n = 86)** | **Agree to participate**<br>**(n = 309)<sup>§</sup>** | **P-value** |
| --- | --- | --- | --- |
| **Age** |  |  | 0.001 |
| <= 24 years old | 17 (19.8) | 122 (39.5) |  |
| > 24 years old | 69 (80.2) | 187 (60.5) |  |
| **Months on PrEP** |  |  | 0.070 |
| <= 3 months | 9 (10.5) | 58 (18.8) |  |
| > 3 months | 77 (89.5) | 251 (81.2) |  |
| **Highest educational level<sup>‡</sup>** |  |  | 0.067 |
| High school or below | 20 (25.0) | 95 (30.7) |  |
| College/ Vocational School | 15 (18.8) | 35 (11.3) |  |
| University | 34 (42.5) | 156 (50.5) |  |
| Post-graduate | 11 (13.8) | 23 (7.4) |  |
| **Employment<sup>‡</sup>** |  |  | 0.047 |
| Unemployed | 4 (5.0) | 40 (12.9) |  |
| Part-time | 14 (17.5) | 71 (23.0) |  |
| Full time | 62 (77.5) | 195 (64.1) |  |
| **Male partner, 12 months<sup>‡</sup>** |  |  | 0.049 |
| No | 1 (1.3) | 0 |  |
| Yes | 79 (98.7) | 309 (100) |  |
| **Male lover, 12 months<sup>‡</sup>** |  |  | 0.466 |
| No | 32 (40.0) | 110 (35.6) |  |
| Yes | 48 (60.0) | 199 (64.4) |  |

<sup>‡</sup>6 observations with missing values refused to participate because they left too quickly so not screening information collected.
<sup>§</sup>1 observation was excluded due to ineligibility after participating in the study.

